# Rare risk variants associate with epigenetic dysregulation in migraine

**DOI:** 10.1101/2021.12.20.21268001

**Authors:** Tanya Ramdal Techlo, Mona Ameri Chalmer, Peter L. Møller, Lisette J. A. Kogelman, Isa A. Olofsson, DBDS Genomic Consortium, Karina Banasik, Mette Nyegaard, Jes Olesen, Thomas Folkmann Hansen

## Abstract

Migraine has a heritability of up to 65%. Genome-wide association studies (GWAS) on migraine have identified 123 risk loci, explaining only 10.6% of migraine heritability. Thus, there is a considerable genetic component not identified with GWAS. Further, the causality of the identified risk loci remains inconclusive. Rare variants contribute to the risk of migraine but GWAS are often underpowered to detect these. Whole genome sequencing is reliable for analyzing rare variants but is not frequently used in large-scale. We assessed if rare variants in the migraine risk loci associated with migraine. We used a large cohort of whole genome sequenced migraine patients (1,040 individuals from 155 families). The findings were replicated in an independent case-control cohort (2,027 migraine patients, 1,650 controls). We found rare variants (minor allele frequency<0.1%) associated with migraine in a Polycomb Response Element in the *ASTN2* locus. The association was independent of the GWAS lead risk variant in the locus. The findings place rare variants as risk factors for migraine. We propose a biological mechanism by which epigenetic regulation by Polycomb Response Elements plays a crucial role in migraine etiology.

## INTRODUCTION

Migraine is a complex neurovascular and genetic disorder which affects 15-20% of the population.^1^ It is characterized by episodes of severe headache, which is typically unilateral and pulsating, accompanied by nausea, vomiting, photophobia and/or phonophobia.^2^ Identifying factors that predispose to migraine is highly relevant. The Global Burden of Disease Study (2019) ranks migraine the highest cause of disability worldwide in young women (aged 15-49) and the second highest cause of disability worldwide among both sexes.^3^ Additionally, migraine has a large socioeconomic impact through treatment costs and loss of productivity estimated at €27 billion in Europe alone.^4^

Migraine has a significant genetic component and is polygenic. There is an increased familial risk and twin studies have shown a heritability of 34%–65%.^5,6^ Multiple, rare variants, each with large effect sizes, may contribute to disease risk together with common variants.^7,8^ The most recent meta-analysis of migraine genome-wide association studies (GWAS) identified 123 migraine risk loci, of which 121 were autosomal. Still, only 10.6% of the total heritability was explained by the GWAS.^9^ GWAS are often underpowered to detect rare variants, as these are imputed and not measured.^10^ Whole genome sequencing (WGS) provides a direct assessment of rare variants. However, it is a technology that is still not frequently used in large-scale and is costly compared to genotyping using arrays.

How the migraine risk loci affect migraine pathogenesis is unknown. Many reside in non-protein coding regions of the DNA and may alter the regulation of gene expression.^11^ Increased risk of migraine has been linked to disruption in gene regulation^12^ and epigenetic regulation.^12–14^ While candidate genes have been identified for increased genetic risk in migraine, the findings have been reported with a high risk of false positive results.^15–18^ This suggests that migraine pathogenesis is affected by mutations outside of protein-coding regions of the DNA, such as in regulatory regions.

We hypothesized that migraine is influenced by rare variants in regulatory regions. We tested if rare variants in the migraine risk loci increase the risk of migraine. We defined blocks of linkage disequilibrium (LD) around the 121 autosomal GWAS lead risk variants using a Danish cohort of 90,312 individuals.^9,19^ Within the LD-blocks, we analyzed rare variants (minor allele frequency (MAF)<0.1%, MAF<1%, and MAF<5%) in genes and regulatory regions using a Sequence Kernel Association Test. For the analysis, we used WGS data from a large cohort of 155 families (*n*_*individuals*_=1,040) with clustering of migraine. The findings were replicated in an independent case-control cohort of migraine patients with no familial history of migraine and controls.

## MATERIALS AND METHODS

### Migraine family cohort

The migraine family cohort consisted of 155 families with a Mendelian-like inheritance pattern of migraine. The participants were recruited from the Danish Headache Center, Rigshospitalet-Glostrup, Denmark, as described elsewhere.^20,21^ All participants were examined by a neurology resident or a senior medical student specifically trained in headache diagnostics. The participants were examined using a semi-structured interview^22,23^ based on the International Classification of Headache Disorders.^2^ The families consisted of 1,040 participants, including 746 individuals diagnosed with migraine and 294 without migraine. The mean number of participants per family was 6.7 and the mean number of migraine patients per family was 4.8. Most of the families were extended beyond the nuclear family. The smallest families consisted of at least two individuals with at least one migraine patient.

### Replication cohort

The replication cohort consisted of 2,027 migraine patients with no familial history of migraine, i.e. unrelated migraine patients, and 1,650 controls. The unrelated migraine patients were recruited and assessed at the Danish Headache Center using the same procedures as described for the family cohort. The controls were recruited for the Danish study of Non-Invasive testing in Coronary Artery Disease (Dan-NICAD) according to procedures described by Nissen *et al*..^24^

### Whole genome sequencing

Blood samples were taken from all participants and genomic DNA was extracted from whole blood. The sequencing was performed on an Illumina NovaSeq 6000 sequencing platform with S4 flow cells. The WGS data was subjected to quality control by deCODE genetics, as described elsewhere.^25^ All samples from both the migraine family cohort and the replication cohort were subjected to the same sequencing and quality control procedures.

### Analytical approach

#### Selecting genomic regions for analysis

We calculated blocks of linkage disequilibrium (LD) around the 121 autosomal migraine risk loci,^9^ using the genomic positions of the GWAS lead risk variants, i.e. the variants with the smallest GWAS *p*-value. For this we used 90,312 participants from the Danish Blood Donor Study (DBDS).^19^ The genomic positions of the GWAS lead risk variants were based on SNP positions from the Database of Single Nucleotide Polymorphisms (dbSNP) build 153 in the GRCh38.p12 (hg38) assembly. The LD-blocks were calculated using a LD-block recognition algorithm proposed by Gabriel *et al*.^26^ with PLINK2.^27^ Here, we assessed variant-pairs within 1000 kilobases of each other with a MAF>5% using a 90% D’ confidence interval threshold >0.70. Subsequently, we annotated genes and regulatory regions within the LD-blocks and analyzed these. If a LD-block partially spanned a gene or regulatory region, we included the entire gene or regulatory region in the analysis. If a LD-block was located in an intergenic region, we extended the genomic region for analysis to include the nearest protein- or RNA-coding gene. No LD-block was generated for 22 loci. In such case, the nearest protein- or RNA-coding gene was included in the analysis. The genomic positions of the regions we analyzed are found in Table S1.

#### Annotating genes and regulatory regions for analysis

The gene transcripts were derived from GENCODE release 36 (basic gene-set only). They included protein-coding genes, non-coding RNA-genes, and pseudogenes. We analyzed the genes in their full length. In addition, we analyzed the coding exons, introns, 5’ untranslated regions (UTR) exons, and 3’ UTR exons by themselves. For regulatory regions, we analyzed promoters, enhancers, CpG islands, insulators, Polycomb Response Elements (PREs), and Transcription Factor Binding Sites (TFBSs). We included promoters from The Eukaryotic Promoter Database (EPD) version 6 and EPD non-coding promoters version 1.^28^ The enhancers were included from the GeneHancer project,^29^ using only enhancers that are supported by multiple studies. The genomic positions of the CpG islands were based on predictions by Gardiner-Garden *et al*..^30^ The insulators and PREs were included from the Broad Institute Chromatin State Segmentation by HMM^31^ from The Encyclopedia of DNA Elements (ENCODE) Consortium^32^ in H1-hESC cells in the GRCh37 (hg19) assembly. The TFBS were included from ENCODE^32^ in the GRCh37 (hg19) assembly. All genomic positions in hg19 were converted to the CRCh38 (hg38) assembly. Table S2 displays the number of genes and regulatory regions that were analyzed in this study.

#### Preparing VCF files for analysis

The VCF files of each participant in the study were merged into one file. Multiallelic sites were converted to a biallelic representation. Subsequently, we annotated genes and regulatory regions to the merged VCF file. We isolated variants, including SNVs, insertions, and deletions, with a MAF<0.1%, MAF<1%, or MAF<5%, using BCFtools.^33^

#### Rare variant association analysis

To test for an association between the rare variants in genes or regulatory regions and migraine, we used the software of Family Sequence Kernel Association Test (F-SKAT).^34^ Age and gender were used as covariates. The age was defined as the age at the time of the interview (time of diagnosis). To avoid association merely given that longer genes and regulatory regions can contain more rare variants, we used modified beta-values.^35^ The beta-values were modified by dividing them with the length of the genes or regulatory regions. The *p*-values were controlled for multiple testing using the Bonferroni method based on the number of genes and regulatory regions tested. A Bonferroni-corrected *p*-value*<*0.05 was considered significant.

To assess if the results were dependent of the GWAS lead risk variants, the number of risk alleles of the variants (0, 1, or 2) were included as covariates in a separate rare variant association analysis with F-SKAT.

#### Replication

The findings were replicated using an independent cohort of unrelated migraine patients and controls. Aggregated data were available on rare variants (MAF<0.1%, MAF<1%, MAF<5%) in the genes and regulatory regions for the controls. We tested if the frequency of the rare variants was increased in the migraine patients compared with the controls, using a Fisher’s Exact Test.^36^ We used the null hypothesis of no association with migraine under a dominant model of penetrance. The resulting *p*-values were controlled for multiple testing using the Bonferroni method. A Bonferroni-corrected *p*-value<0.05 was considered significant.

#### Characterization of findings

To explore the effect of the rare variants of the replicated results, we used Ensembl Variant Effect Predictor.^37^ To discover the target genes of the replicated results, we used *cis*-eQTLs data from the Genotype-Tissue Expression (GTEx) version 8.0 dataset (dbGaP Accession phs000424.v8.p2 release).

#### Ethics

Oral and written consent was obtained from all participants. All procedures performed in studies involving human participants were in accordance with the ethical standards of the National Committee on Health Research Ethics (file no. H-2-2010-122) and with the 1964 Helsinki declaration and its later amendments. The study was approved by the data-protection agency (01080/GLO-2010-10).

## RESULTS

### Rare variants in regulatory regions associate with migraine

We found an association between migraine and rare variants with a MAF<0.1% in 7 regulatory regions and an intron of the *MACF1* gene (see Table 1 for genomic regions and *p*-values). No association was found for rare variants with a MAF<1% or a MAF<5% nor in coding regions (full gene transcripts, coding exons, 5’ UTR exons, 3’ UTR exons). In line with what is expected when analyzing rare variants for polygenic traits,^9,38,39^ we found that the distribution of the test statistics deviated from the null with inflated genomic inflation factors (λ=1.89, λ=1.63, and λ=1.68) (Figure S1).

**Table 1.**
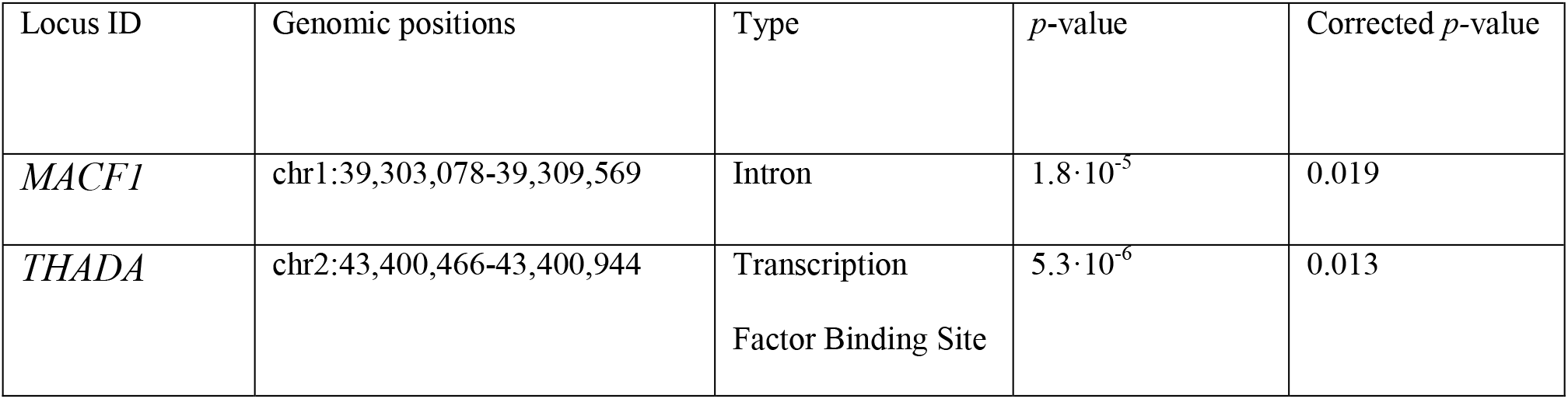
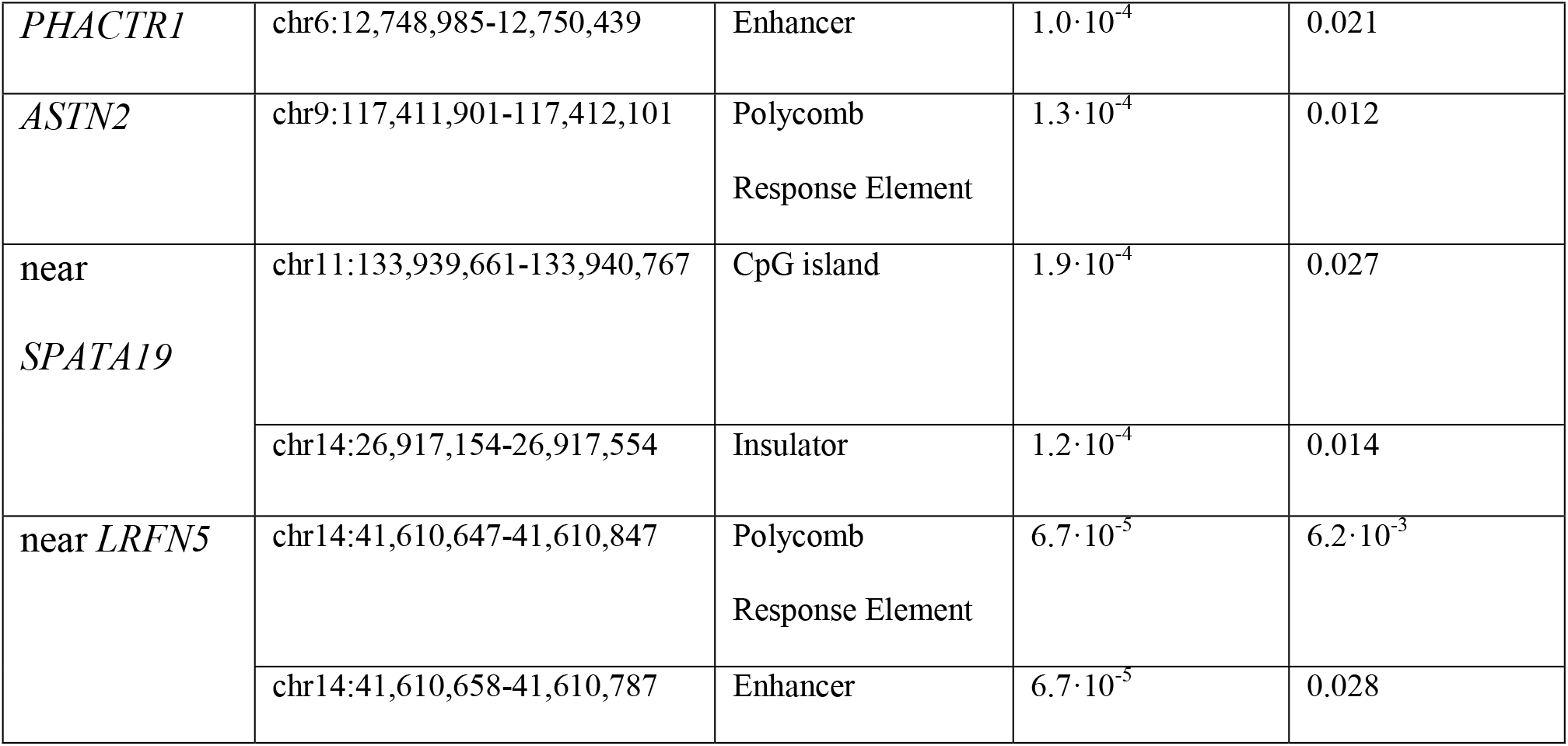
Summary of the results obtained from the rare variant association analysis. The results from the rare variant association analysis together with the locus ID, as given in the meta-analysis of migraine GWAS by Hautakangas *et al*.^9^, the genomic positions in GRCh38.p12 (hg38), and type of genomic feature.

Subsequently, we replicated the Polycomb Response Element (PRE) located at chr9:117,411,901-117,412,101 in the *ASTN2* locus (*p*=1.1·10^−15^) using an independent case-control cohort, henceforth referred to as the replicated PRE.

We found that the findings were independent of the GWAS lead risk variants, with unaffected results (*p*=1.3·10^−4^ in the original analysis (see Table 1) and *p*=1.2·10^−4^ with the allele count of the risk variants as covariates).

### Rare variants in PRE map to CTCF binding sites

The replicated PRE was located in the first intron of the *ASTN2* gene and overlapped the TFBS of CTCF, SIN3A, ZNF444, GATA2, RCOR1, and MYC (Figure 1). There were 12 rare variants with a MAF<0.1% located in the replicated PRE. These rare variants consisted of 1 SNV, 9 deletions, and 2 insertions (Table 2). We found that all rare variants mapped to CTCF binding sites (Table 2) using the Ensembl Variant Effect Predictor (VEP).

**Figure 1.**
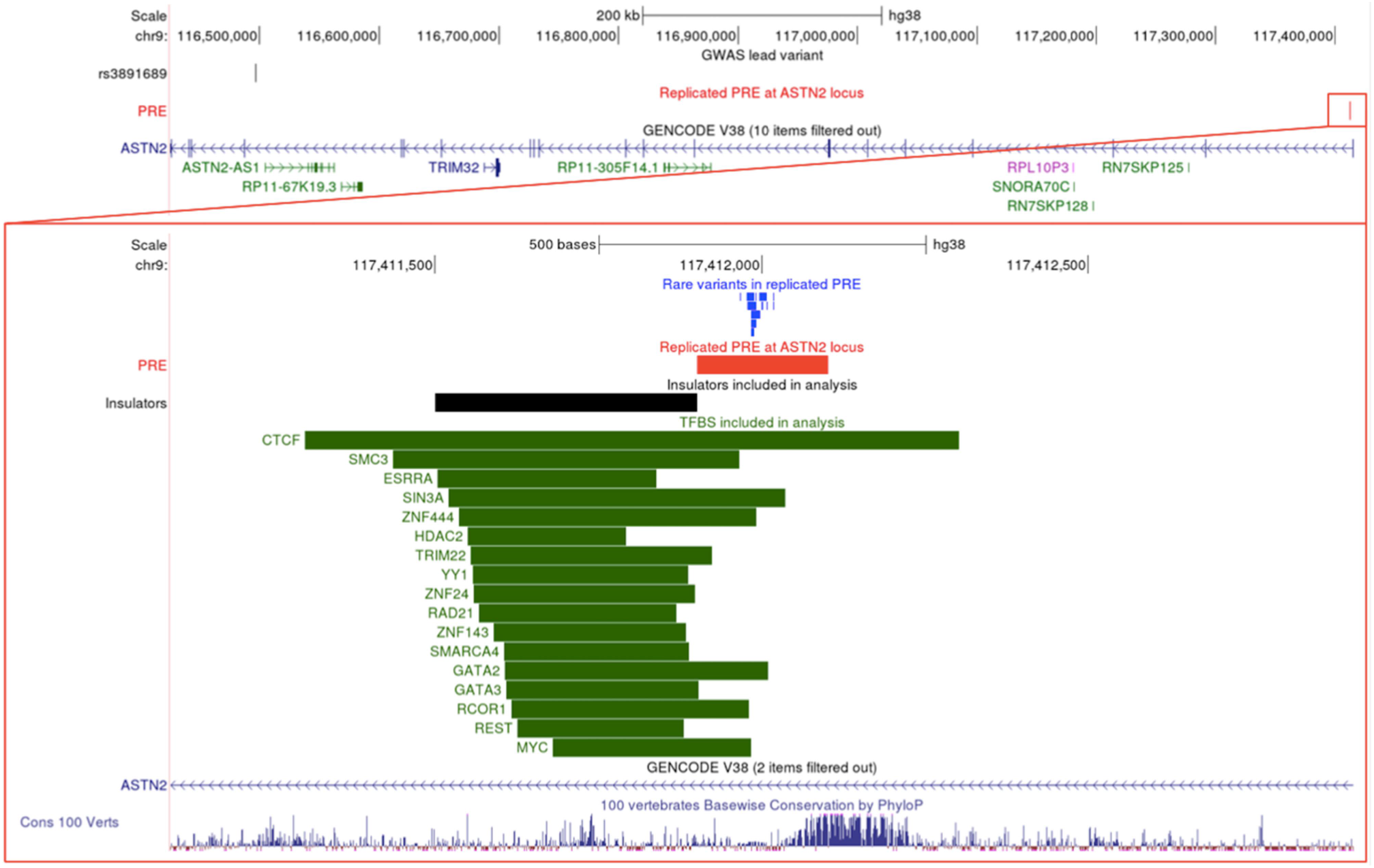
Overview of the genomic region around the *ASTN2* migraine risk locus. Top: The genomic region that was included for analysis (see Table S1 for genomic positions). Displayed is the GWAS lead risk variant (rs3891689), the replicated Polycomb Response Element, and the genes included for analysis. **Bottom:**A close-up of the replicated Polycomb Response Element and its surrounding genomic region. Displayed are the rare variants with a MAF<0.1%, that associated with migraine, the replicated Polycomb Response Element, near-by regulatory regions (an insulator and 17 transcription factor binding sites), the first intron of the *ASTN2* gene, and the evolutionary conservation across 100 vertebrates.

**Table 2.** Information on the rare variants in the replicated Polycomb Response Element. The table presents the genomic positions in GRCh38.p12 (hg38), the reference allele, the alternative allele, the variant type, the rs number, and the Ensembl regulatory feature of the rare variants (MAF<0.1%) in the replicated Polycomb Response Element.

| Genomic position | Reference allele | Alternative allele | Variant type | rs number | Ensembl regulatory feature |
| --- | --- | --- | --- | --- | --- |
| chr9:117411967-117411967 | A | C | SNV | rs866789329 | CTCF binding site |
| chr9:117411977-117411988 | CCACCCCTACCA | C | Deletion |  | CTCF binding site |
| chr9:117411978-117411991 | CACCCCTACCACCA | C | Deletion | rs158802387<br>2 | CTCF binding site |
| chr9:117411983-117411997 | CTACCA | C | Deletion | rs144647713<br>7 | CTCF binding site |
| chr9:117411983-117411991 | CTACCACCA | C | Deletion |  | CTCF binding site |
| chr9:117411983-117411988 | CTACCACCACCCCC<br>A | C | Deletion |  | CTCF binding site |
| chr9:117411990-117411991 | CA | C | Deletion |  | CTCF binding site |
| chr9:117411996-117412007 | CACCTCACCCCT | C | Deletion |  | CTCF binding site |
| chr9:117411999-117412002 | CTCA | C | Deletion | rs138331219<br>5 | CTCF binding site |
| chr9:117412007-117412008 | TC | T | Deletion | rs145255224 | CTCF binding site |
| chr9:117412018-117412018 | A | ACCCCCCCCCC | Insertion |  | CTCF binding site |
| chr9:117412018-117412018 | A | ACCCCCCCCCC<br>C | Insertion |  | CTCF binding site |

### Possible regulatory targets unidentified

Finally, we assessed the possible regulatory targets of the replicated PRE. We found no significant *cis*-eQTLs in the PRE using the GTEx database for the identification of regulatory targets.

## DISCUSSION

Our study is, to our knowledge, the first to assess if rare variants in 121 autosomal migraine risk loci increase the risk of migraine. We assessed if rare variants in genes and regulatory regions associated with migraine, using WGS data from families with clustering of migraine. We replicated a Polycomb Response Element (PRE) in the *ASTN2* locus using an independent case-control cohort.

### Regulatory regions harbor rare variants associated with migraine

We found an association between migraine and rare variants with a MAF<0.1% in 7 regulatory regions and an intron of the *MACF1* gene. Our results indicate that migraine can be associated with rare variants in regulatory regions, since introns are likely to harbor elements with a regulatory function.^40^ We replicated a PRE in the *ASTN2* locus using an independent case-control cohort. Interestingly, we have previously reported a rare variant association with migraine in a PRE in the same locus, using a different study design.^12^ Importantly, we show that the effect of the replicated PRE was independent of the GWAS lead risk variant. Thus, our results show that migraine GWAS loci also contain independent rare risk variants.

### Disruption in epigenetic regulation as a possible biological mechanism for migraine

The function of mammalian PREs is still largely unknown.^41^ Mammalian PREs are likely capable of binding Polycomb Group (PcG) proteins as a part of transcriptional silencing during cell division, where the silencing is maintained in subsequent daughter cells.^41–43^ Associated with the silencing is remodeling of the chromatin.^44,45^ Possibly, the rare variants we find associated with migraine in the replicated PRE can cause a disruption in the binding of PcG proteins and chromatin remodeling. The disruption may lead to transcriptional silencing not being maintained after cell division. This ultimately leads to an altered gene expression (Figure 2). Epigenetic mechanisms have been linked to brain plasticity in the transition from episodic to chronic migraine.^13,14^ However, we are, to our knowledge, the first to report an indication of an actual biological mechanism for migraine, where chromatin remodeling is disrupted by rare variants in PREs.

**Figure 2.**
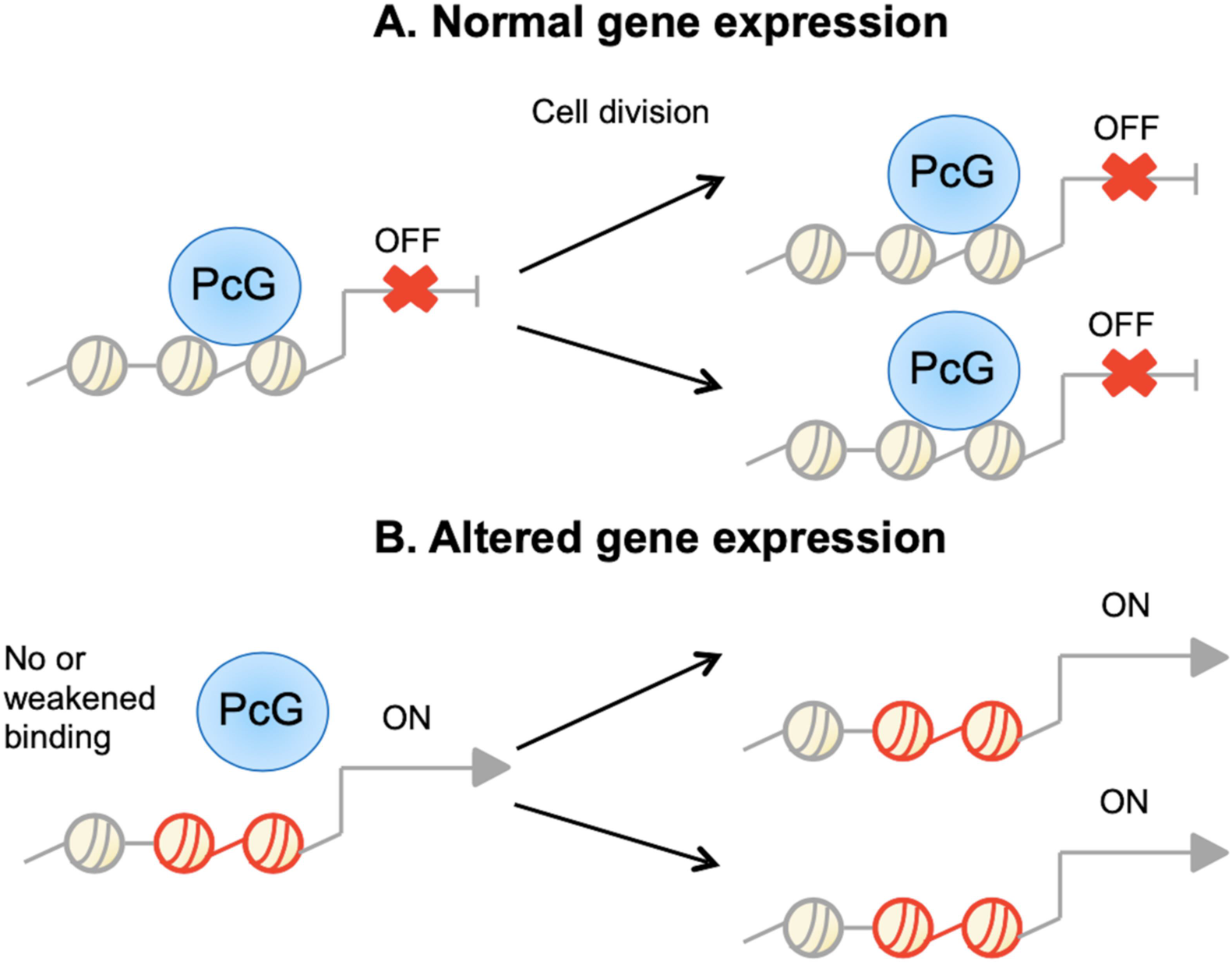
Schematic overview of transcriptional silencing by PcG proteins in absence or presence of rare variants in the DNA. **A**. The recruitment of PcG proteins to the DNA results in transcriptional silencing that is maintained after cell division. **B**. A disruption of PcG binding, e.g. by rare variants, results in no binding or a weakened binding of the PcG proteins. A disrupted binding leads to an altered gene expression where gene silencing is disturbed. The altered gene expression is maintained after cell division.

We found that the rare mutations in the replicated PRE were situated in binding sites of the transcription factor CTCF. CTCF is a transcriptional repressor, with many functions including regulation of gene expression through chromatin modifications.^46^ Again, our findings indicate a link to a disruption in chromatin remodeling as a biological mechanism for migraine.

### *ASTN2* as a possible regulatory target gene

We were not able to identify regulatory targets of the replicated PRE based on significant *cis*-eQTLs. Thus, the target gene could be assigned to the nearest gene, as done by Ringrose *et al*..^47^ The nearest gene is *ASTN2*, of which the PRE is overlapping. The *ASTN2* gene encodes the Astrotactin 2 protein. The Astrotactin 2 protein is expressed in the brain and is suggested to play a part in neuronal migration. Thus, our findings support the hypothesis of migraine being a neuronal disorder.^48^

### Implications for genetic studies on migraine

The most recent meta-analysis of migraine GWAS only explains 10.6% of the total heritability of migraine.^9^ GWAS are often underpowered to detect rare variants with low-penetrant alleles.^10^ We found that migraine associated with very rare variants, i.e. variants with a MAF<0.1%, in the migraine risk loci. Our results indicate that a part of the unexplained heritability of migraine could arise from rare variants not detected by GWAS. Also, our results suggest that the unexplained heritability could be hidden in epigenetics. For discovering the unexplained heritability of migraine, future studies focusing on the identification of heritable epigenetic marks for migraine can be highly relevant.

## CONCLUSION

We found that rare variants (MAF<0.1%) in a Polycomb Response Element in the *ASTN2* locus associated with migraine. The association was independent of the original, common variant in the locus. Thus, migraine GWAS loci also contain independent rare risk variants. We propose a biological mechanism for migraine by which chromatin remodeling is disrupted by rare variants in Polycomb Response Elements.

## Data Availability

All data produced in the present study are available upon reasonable request to the authors

## ABBREVIATIONS

GWAS: Genome-Wide Association Study
LD: Linkage disequilibrium
MAF: Minor allele frequency
PcG: Polycomb Group protein
PRE: Polycomb Response Element
TFBS: Transcription factor binding site
UTR: Untranslated region
WGS: Whole genome sequencing

## FUNDING

The study was funded by a grant from Candys Foundation “CEHEAD” and a grant from Jascha Fonden (web site: https://jaschafonden.dk/). The funders had no role in study design, data collection and analysis, decision to publish, or preparation of the manuscript.

## COMPETING INTERESTS

The authors have declared that no competing interests exist.

## DATA AVAILABILITY

The data supporting the conclusions of this article are unavailable, as they contain information that may be used for identifying study participants. However, summary data supporting the conclusions of this article are available on request to the corresponding author of this article.

## ACKNOWLEDGEMENTS

We thank deCODE genetics for whole genome sequencing and quality-controlling our samples. We thank the Danish blood donors for their valuable participation in the Danish Blood Donor Study as well as the staff at the blood centers for making this study possible. Finally, we thank all of the staff and participating patients at the Danish Headache Center for their willing assistance.

## CONSORTIA

Members list of the DBDS Genomic Consortium:

Steffen Andersen^1^, Karina Banasik^2^, Søren Brunak^2^, Kristoffer Burgdorf^3^, Christian Erikstrup^4^, Thomas Folkmann Hansen^5^, Henrik Hjalgrim^6^, Gregor Jemec^7^, Poul Jennum^8^, Per Ingemar Johansson^3^, Kasper Rene Nielsen^9^, Mette Nyegaard^10^, Mie Topholm Bruun^11^, Ole Birger Pedersen^12^, Susan Mikkelsen^4^, Khoa Manh Dinh^4^, Erik Sørensen^3^, Henrik Ullum^3^, Sisse Ostrowski^3^, Thomas Werge^13^, Daniel Gudbjartsson^14^, Kari Stefansson^14^, Hreinn Stefánsson^14^, Unnur Þorsteinsdóttir^14^, Margit Anita Hørup Larsen^3^, Maria Didriksen^3^, Susanne Sækmose^12^.

^1^ Department of Finance, Copenhagen Business School, Copenhagen, Denmark.

^2^ Novo Nordisk Foundation Center for Protein Research, Faculty of Health and Medical Sciences, University of Copenhagen, Copenhagen, Denmark

^3^ Department of Clinical Immunology, Copenhagen University Hospital, Copenhagen, Denmark

^4^ Department of Clinical Immunology, Aarhus University Hospital, Aarhus, Denmark

^5^ Danish Headache Center, Department of Neurology, Copenhagen University Hospital, Glostrup, Denmark

^6^ Department of Epidemiology Research, Statens Serum Institut, Copenhagen, Denmark

^7^ Department of Clinical Medicine, Sealand University Hospital, Roskilde, Denmark

^8^ Department of Clinical Neurophysiology, University of Copenhagen, Copenhagen, Denmark

^9^ Department of Clinical Immunology, Aalborg University Hospital, Aalborg, Denmark

^10^ Department of Biomedicine, Aarhus University, Aarhus, Denmark

^11^ Department of Clinical Immunology, Odense University Hospital, Odense, Denmark

^12^ Department of Clinical Immunology, Næstved Hospital, Næstved, Denmark

^13^ Institute of Biological Psychiatry, Mental Health Centre Sct. Hans, Copenhagen University Hospital, Roskilde, Denmark

^14^ deCODE genetics, Reykjavik, Iceland

## Notes

### Competing Interest Statement

The authors have declared no competing interest.

### Funding Statement

Candy's Foundation (CEHEAD)
Jascha Fonden

### Author Declarations

Danish Scientific Ethical Committee of Region H gave ethical approacl of this work, no. H-2-2010-122. The Danish Data Protection Agency of region H as approaved the project, 01080/GLO-2010-10.

